# Survey Design and Geospatial Analysis using Data on Baseline Prevalence, Environmental Risk-Factors and Treatment History Drastically Reduces the Cost of STH Impact Surveys

**DOI:** 10.1101/2023.01.03.23284146

**Authors:** Claudio Fronterrè, Olatunji Johnson, Emanuele Giorgi, Antonio Montresor, Peter J Diggle

## Abstract

Soil-transmitted helminth (STH) infections are among the most common neglected tropical diseases worldwide, causing high morbidity and mortality rates in endemic areas. STH impact assessment surveys provide information on the changing epidemiological situation so that control programme managers can adapt the frequency of any continuing preventive chemotherapy (PC). The cost associated with conducting these surveys is an important factor, especially in developing countries with severely limited financial resources. Using three case studies based on historical data on baseline prevalence, environmental risk-factors and treatment history from Kenya, Sierra Leone and Zimbabwe, we show that a model-based geostatistics approach drastically reduces the cost of STH impact surveys by comparison with a survey conducted according to the current WHO guidelines, whilst maintaining the same overall accuracy. The framework that we develop is disease-agnostic and could easily be repurposed for use with other NTDs.

## 1 Introduction

Soil-transmitted helminth (STH) infections are the most widespread of the neglected tropical diseases, primarily affecting marginalized populations in low- and middle-income countries; More than one billion people are currently infected with one or more STH species; these include *Ascaris lumbricoides, Trichuris trichiura, Necator americanus* and *Ancylostoma duodenale* [1] The World Health Organization (WHO) recommends an integrated approach to the control of morbidity due to STH, which includes access to appropriate sanitation, hygiene education, and preventive chemotherapy (PC). [2]

In the last 10 years PC coverage has significantly scaled up, to reach approximately 60% of the children in need by 2019[3]. This has resulted in a progressive decline of prevalence and intensity of STH infections globally [4]. Managers of control programme in endemic countries need periodically to conduct an STH Impact Assessment Survey (IAS) in order to adapt the frequency of PC to the changing epidemiological situation. An IAS is typically conducted after 5 years of PC interventions with Albendazole (ALB) or mebendazole (MBZ). Its aim is to estimate current prevalence and so determine whether, and if so at what frequency, PC needs to continue. The WHO’s criterion for suspension of PC is that local prevalence of infection with any STH is less than 2% [5].

The cost associated with conducting an IAS is a challenge to control programs, especially in developing countries with scarce technical and financial resources. Efforts have been made to reduce the cost by conducting school-based surveys, which focus on the target population of school-aged children (SAC), to estimate prevalence at implementation unit (IU) level, and use this information to decide the frequency of continuing PC.

We have previously shown that model-based geostatistical methods can deliver an order-of-magnitude improvement in the precision of prevalence surveys by comparison with traditional survey methods [6, 7]. Our aim in this paper is to extend this methodology to the efficient design and analysis of an IAS.

Traditionally, an IAS is conducted with the same methodology used for an earlier baseline survey, but is analysed without reference to baseline data. We shall show that taking advantage of the large amount of data collected at baseline and during program implementation makes it possible to reduce drastically the sample size required, and therefore the cost of an IAS, without degrading the precision of the survey.

Most STH-endemic countries have conducted a baseline survey (i.e. pre-MDA). We shall assume that prevalence data are available from a baseline prevalence survey conducted in each of a number of geolocated schools within the region of interest, *A*. Our goal is to use this information in combination with environmental risk factor data and PC history (i.e. frequency and coverage) to develop a more cost-efficient IAS at any time in the future.

In summary, our proposal is: firstly, to fit a geospatial statistical model [8] to the data available at baseline and use the fitted model to predict baseline prevalence throughout *A*; secondly, to project the prevalence surface forward to the time of the proposed impact survey, incorporating whatever additional information, for example number, frequency and coverage of PC or other interventions post-baseline, has accrued in the meantime; thirdly, to create different scenarios of sampling design using the projected prevalence surface; finally, to compare different designs according to a specified performance measure.

In what follows, we applied this method to baseline data collected in Kenya, Zimbabwe and Sierra Leone. In each of these countries both baseline and IAS data are available, which allows us to evaluate empirically the performance of our proposed methodology.

## 2 Designing an impact assessment survey

### 2.1 Ingredients

The *minimal information-set* needed to design an impact survey over a designated geographical area *A* consists of:

1. baseline prevalence data collected before any PC;
2. PC treatment coverage history since baseline, usually available at *implementation unit* (IU) level;
3. a model for the transition from baseline to impact that provides a rough estimate of the expected prevalence at impact conditional on the frequency and coverage of PC;
4. the set of school or community geolocations that could be considered for inclusion in the IAS or, failing this, the approximate number of schools or communities in each IU. A raster image of population density throughout *A* can be used to impute approximate geolocations of schools when their exact locations are not available beforehand.

Additionally, raster images of environmental risk-factors may enable more precise predictions at negligible additional cost, depending on the strength of their association with local prevalence.

An *impact survey design* consists of a set of *m* sample locations (schools/communities) to be sampled from a larger set of potential sample locations and the number, *n*, of individuals to be sampled at each location. We will assume that at any location, the sampled individuals are selected at random from the population at risk, whilst the locations will be selected according to a *spatially regulated design*, defined as a random selection subject to the constraint that no two sampled locations can be separated by less than a designated distance *δ*. The random selection can either be uniform (referred to as non-stratified) or non-uniform (referred to as stratified). To avoid subjective bias, this requires the specification of an *inclusion function, if* (*x*), such that the probability of a location *x* being included in the design is proportional to *if* (*x*); for example, it might be desirable deliberately to over-sample areas whose prevalence is thought be high. Hence, a design construction, 𝒟 is defined by four quantities and is denoted by 𝒟 = 𝒟 (*m, n, if, δ*). Any such construction can be applied to the whole of *A* or separately within each of a set of sub-areas or *strata* that partition *A*. For example, we might choose to stratify by district and constrain the design to sample a prescribed number of locations in each district. Here, we assume that 𝒟 applies to the whole of *A*.

To evaluate a proposed design construction, and hence to optimise the design subject to any practical constraints, we need to define a *performance measure*. Ideally, this would include consideration of the relative costs and benefits associated with correct and incorrect decisions regarding continuation of PC, and a limit on the actual cost of conducting the survey. The flow diagram reported in Figure 1 shows the schematic of the whole process.

**Figure 1:**
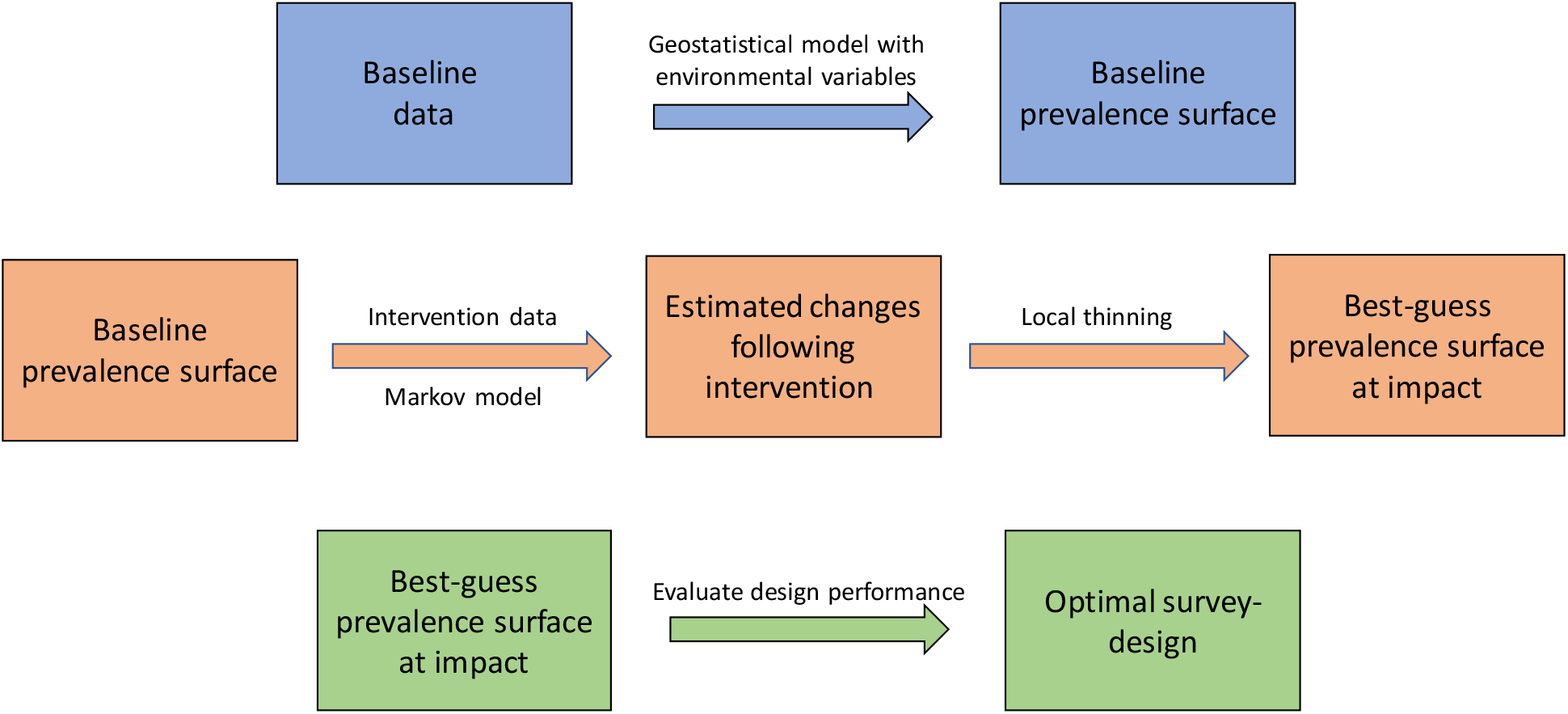
Schematic representation of the stages needed for optimising the survey design of STH impact assessment surveys.

### 2.2 Predicting baseline surface prevalence

Model-based geostatistical (MBG) methods enable prediction of prevalence at both surveyed and unsurveyed locations. For this reason, they have been widely used for prevalence mapping in a range of public and global health applications, but especially in low resource settings where disease registries do not exist. In these circumstances, prevalence mapping must rely on data collected from surveys of disease prevalence taken in a sample of the schools/communities at risk within the region of interest, supplemented by remotely sensed images that measure, or can act as proxies for, environmental risk factors. MBG methods have been used for prevalence mapping of loaiasis [9], onchocerciasis and [10] malaria [11], among others. In settings where geographical variation in prevalence exhibits spatial correlation, MBG methods can give substantial gains in efficiency over traditional methods [6].

To infer the true baseline prevalence surface, *P*_0_(*x*), we require baseline prevalence surveys. These data consist of a set of locations, usually schools or communities, at each of which a pre-specified number of individuals is tested for STH prior to any PC. If data are also available as raster images of demographic and environmental variables that can potentially explain part of the variation in prevalence, such as temperature, precipitation, elevation, Normalized difference vegetation index (NDVI) and soil moisture, their incorporation into the model as covariates can improve predictive precision at the cost of making assumptions about the form of their relationship with prevalence.

We fit a geostatistical model to the data to generate the baseline prevalence surface, as follows. The sampling distribution of the number, *Y*_*i*_, of individuals who test positive at sampling location *x*_*i*_ is binomial, with number of trials *n*_*i*_ and probability of positive outcome *P*_0_(*x*_*i*_). The variation in *P*_0_(*x*_*i*_) is modelled using a combination of demographic and environmental covariate effects *d*(*x*), unexplained residual spatial variation, *S*(*x*), and unstructured random effects, *Z*_*i*_. The resulting geostatistical model for prevalence data is a generalized linear mixed model,

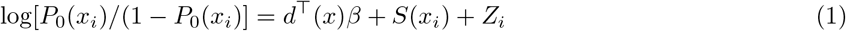

where *β* is the corresponding regression coefficient associated with *d*(*x*). The geostatistical model is fitted separately to the three STH species to allow for different levels of spatial correlation and for species-specific covariate effects.

### 2.3 Estimating changes following intervention

To project the prevalence surface forward to the time of the proposed impact survey, we follow a two-stage procedure. Firstly, for each species, we use a *multi-state Markov model* to estimate the transition probabilities between endemicity classes as a function of PC history. Secondly, we perform a *local thinning* of the predicted baseline prevalence surface to generate our best guess of the impact prevalence surface.

#### 2.3.1 Multi-state Markov model for predicting the endemicity class

To fit the model, we require at least the following data: prevalence data from two time points (years), *t* and *t* + *k*; covariate data that affect the probability distribution of prevalence at time *t* + *k* given prevalence at time *t*. We then classify the observed prevalence into *J* = 5 endemicity classes or states: (*c*_1_ = [0 − 2%), *c*_2_ = [2% − 10%), *c*_3_ = [10% − 20%), *c*_4_ = [20% − 50%), *c*_5_ = [50% − 100%]) and estimate the Markov transition probability (MTP) matrix, *π*, whose elements *π*_*ij*_ are the probabilities of transitioning from endemicity state *i* to endemicity state *j*, by fitting a separate multinomial logistic model conditional on each baseline endemicity state. Specifically, we assume that the key epidemiological predictor of future prevalence is the PC coverage history. Therefore, we include as a covariate the number of *effective PC rounds*, defined as the number of rounds that are reported to have achieved epidemiological coverage of at least 75% at IU-level. Hence, the multinomial logistic model for the transition probability *π*_*ij*_ is

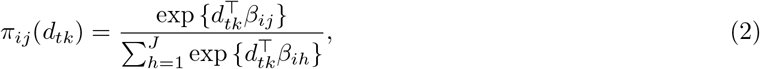

where *d*_*tk*_ is the number of effective PC rounds between times *t* and *t* + *k*.

To predict the transition probability at any future time point *t* + *s*, for *s* > *k*, we use the estimated regression coefficients 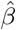 and replace *d*_*tk*_ in equation (2) by *d*_*ts*_, the number of effective PC rounds from time *t* to time *t* + *s*.

The final step is to predict the endemicity class of the future impact survey for each IU. Let *ω*_*t*_ = {*ω*_*t*_(*c*) : *c* = 1, …, 5} denote the probability distribution of the IU-wide population-weighted prevalence over the five endemicity states at baseline. We calculate the probability vector *ω*_*t*+*s*_ at time *t* + *s* as

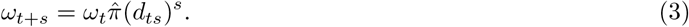

with 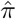 the maximum likelihood estimate of the MTP matrix *π*. Then from *ω*_*t*+*s*_, we can estimate the endemicity class *ĉ* at time *t* + *s*, as the class corresponding to the largest element of the probability vector, *ω*_*t*+*s*_. To reflect the fact that the three STH species react differently to PC treatment we estimate species-specific transition probabilities. We also used prevalence data from Kenya collected at baseline and after three years of treatment to estimate the transition probabilities and then applied the same transition probabilities to Sierra Leone and Zimbabwe.

#### 2.3.2 Local thinning to generate the impact surface

To obtain an estimate of the prevalence surface at impact, *P*_1_(*x*), we scale-down the predicted baseline prevalence surface for each species at IU level according to the endemicity class estimated by the Markov model and then aggregate across species, assuming independence. For each IU, we calculate the log-odds surface at baseline, 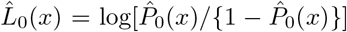, and the projected log-odds surface at impact, 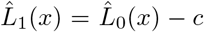, where *c* is a constant such that the population-weighted mean prevalence at IU level is equal to the lower bound of the estimated endemicity class. Converting 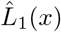 back to the prevalence scale gives us an estimate 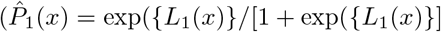, that we use as our best guess for the true prevalence surface *P*_1_(*x*) in a simulation-based evaluation of the performance of different survey designs.

### 2.4 Design the survey

We can test a set of candidate survey design constructions *D*(*m, n, if, δ*) by simulating prevalence data from our best guess of the prevalence surface at impact, 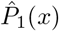. In our application the candidate locations that constitute the sampling units are primary schools. If a database of geolocated schools is not available we simulate it according to census data and the underlying population density. We tested 40 candidate survey designs obtained by varying the four key design elements as described in Section 3.

### 2.5 Evaluating the performance

We evaluated the performance of the proposed IAS design in two ways: reduction of sample size; and in accuracy of the indicated treatment decision. The general goals of an IAS are to evaluate the current burden of the disease after 5 to 6 years of PC, and to classify each IU, typically a district, or sub-district, into one of five endemicity classes. Each endemicity class determines a treatment decision for the IU in question. For each design, we compared the classification of each IU given by the proposed geostatistical approach with the one given by the classical approach, by which we mean the current practice of sampling schools and children at random without clear guidelines on how to choose the sample size. This is also coupled with a decision algorithm that relies on the observed empirical prevalence rather than the output of a model that exploits the spatial correlation present in the data.

Within our MBG framework, we can assign to each IU a probability distribution, *{p*_*IU*_ (*c*) : *c* = 1, …, 5*}* of the population-weighted prevalence over the five pre-defined classes. One way to assign a single predicted class *ĉ* is as the mode of the distribution, i.e. the value of *c* corresponding to the largest of the five probabilities *p*_*IU*_ (*c*). We can then compare the predicted class with the true class and evaluate the design according to an agreed set of performance measures.

## 3 Results

We have applied the methodology described in this paper to three different geographical settings, Kenya, Sierra Leone and Zimbabwe. First, we used the data from Kenya to estimate the effect of the number of effective PC rounds on the change in endemicity state as shown in equation 2. We used the Kenya data for this task because of its longitudinal structure whereby the same set of schools was repeatedly surveyed at three different time points, 2012, 2015 and 2017. We estimated transition probabilities from the 2012 and 2015 survey data, and used the resulting estimates in our subsequent evaluations of the performance of each candidate design, using the results from actual impact surveys already collected in each of the three countries as a gold-standard.

Figure 2 show the predicted surfaces of the prevalence at baseline (left panel), projected impact (middle panel) and actual impact (right panel), for Kenya, Sierra Leone and Zimbabwe, respectively. The baseline and the actual impact prevalence surface were obtained from an MBG model that incorporates a set of demographic and environmental variables, while the projected impact surface was obtained from the multi-state Markov and local thinning approach described in section 2.3. For the Kenya surface, the projected prevalence surface is quite similar to the actual surface, meaning that the PC intervention history wasvery effective in projecting the prevalence surface from baseline to impact. For Sierra Leone and Zimbabwe, we found higher discrepancies between the projected prevalence surface and the actual prevalence surface. One reason for this is that Kenya data was used to estimate the Markov transition parameters, and the endemicity level of Kenya is quite different from that of the other two countries; prevalence is much lower in Zimbabwe, and much higher in Sierra Leone.

**Figure 2:**
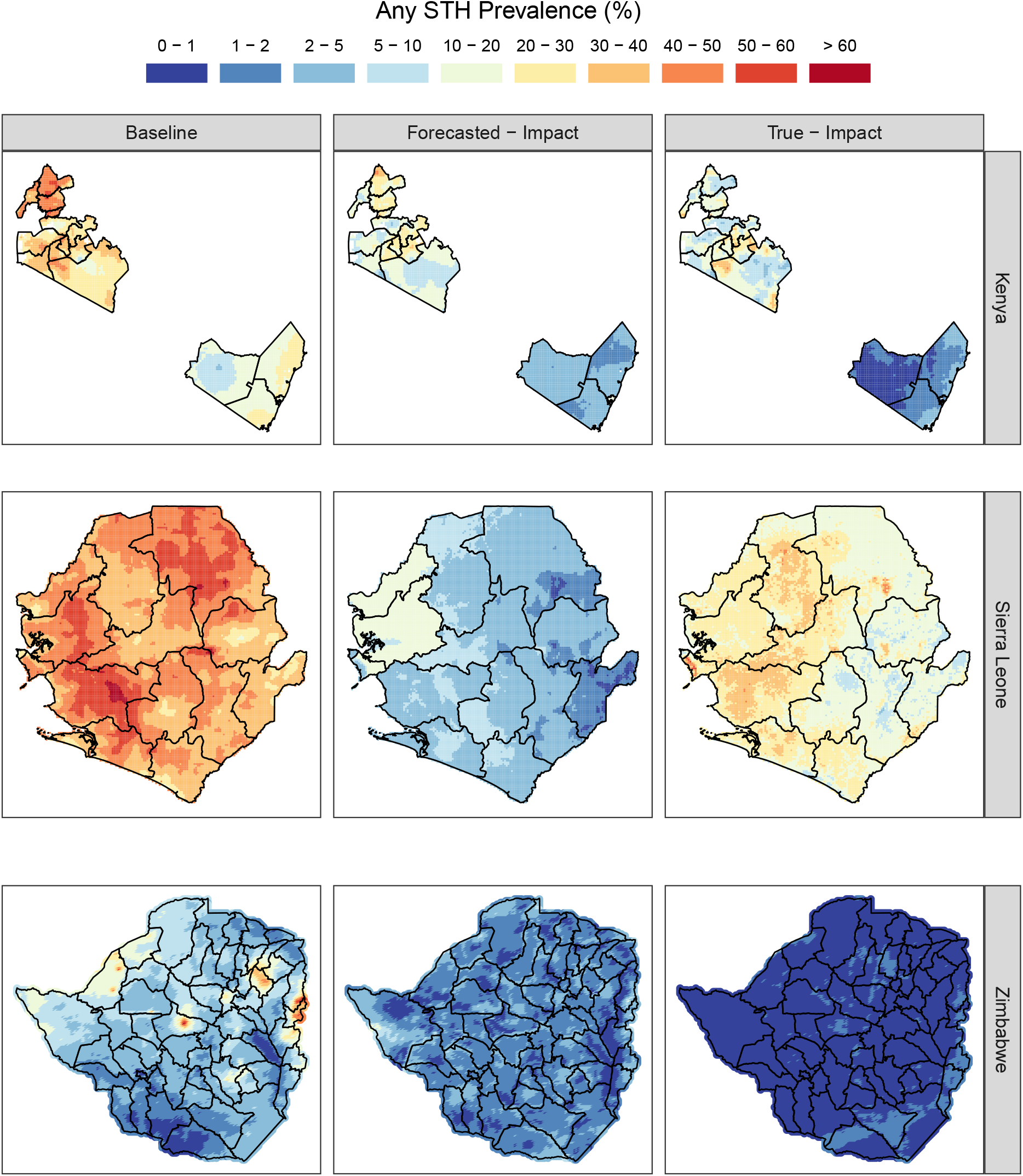
Predicted prevalence surfaces at baseline (left panel), projected prevalence surface at IA (middle panel) and the true prevalence surface at IA (right panel) of any STH in Kenya, Sierra Leone and Zimbabwe

Figure 3 shows, for each combination of schools and children sampled, the approximate cost of the the survey based on unit cost estimates of 200$ for sampling a school and 10$ to test a child, giving a total survey cost,

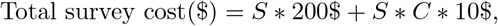

where *S* is the number of schools sampled and *C* is the number of children tested in each school. The two dots reported in each panel of Figure 3 indicate pairs of surveys, one conducted with the classical and one with the MBG approach, that have the same accuracy. The MBG approach achieves the same level of accuracy as the classical approach but with a substantially smaller sample size. The average decrease in survey cost is 80% with reductions for Zimbabwe, Kenya and Siera Leone of 91%, 86% and 62%, respectively.

**Figure 3:**
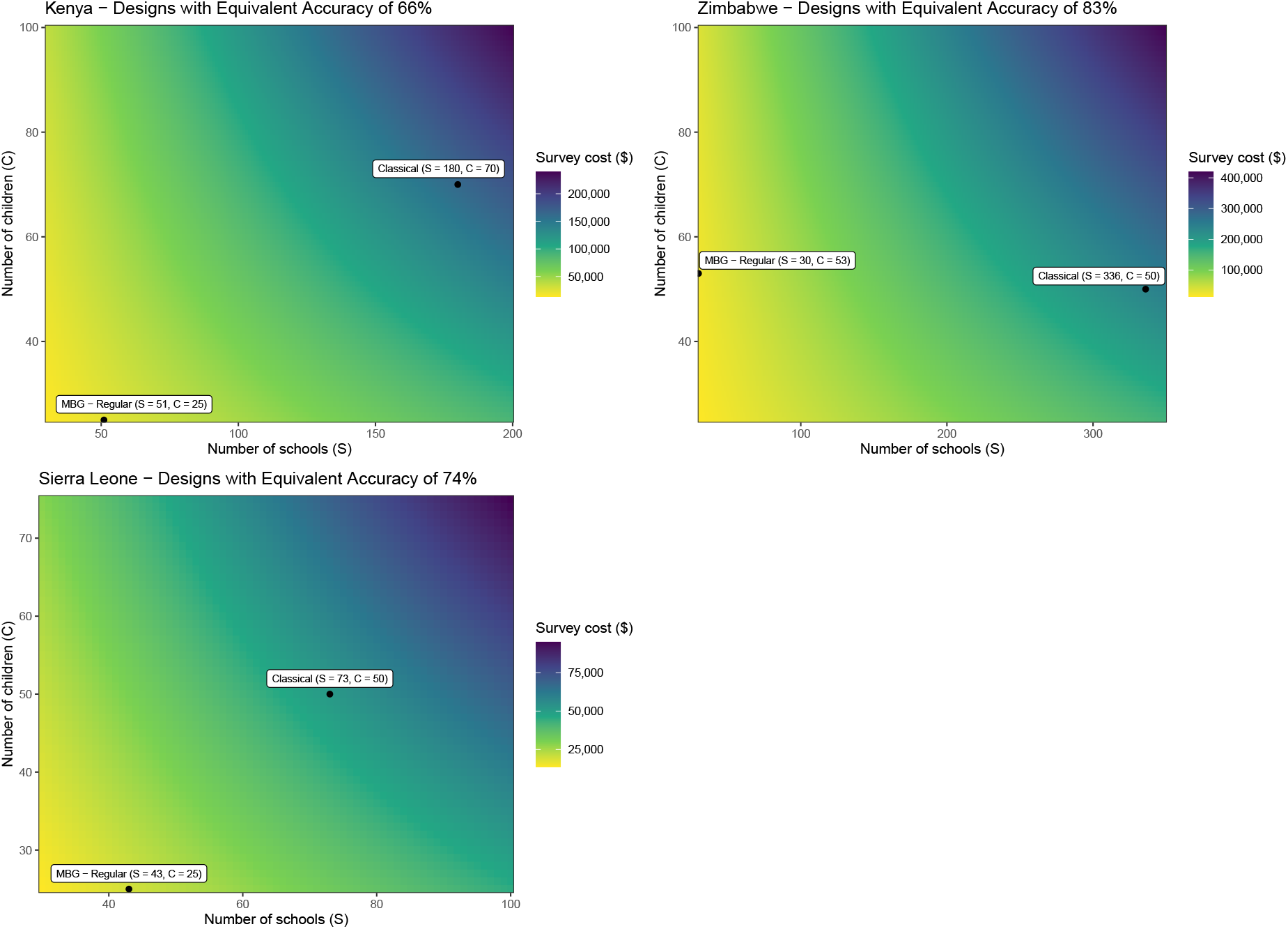
Simulation results for Kenya, Zimbabwe and Sierra Leone.

## 4 Discussion

We have demonstrated that the MBG approach, incorporating data on baseline prevalence, environmental risk-factors and treatment history, can deliver impact assessments with substantially higher accuracy at substantially lower cost than the classical approach.

This methodology, if applied in the three country examples considered, could have resulted in a drastic reduction in the number of schools to be visited and the number of children to be investigated in each school, resulting in a survey cost in the order of one-fifth the cost of a classical survey conducted according to current WHO guidelines whilst maintaining the same overall accuracy. These savings are especially valuable in context of the limited resources available for control of STH in most affected countries. The MBG approach also delivers finely resolved maps of prevalence at impact that could form an important element in promoting self-sustainability of all aspects of STH control in endemic countries, for example by identifying residual sub-IU-level hotspots that wold be prime candidates for further intervention or post-elimination surveillance.

We are developing a manual to help countries to apply the MBG methodology, and a WEB application to facilitate its practical implementation. Although these resources are currently being developed specifically for STH, the framework is disease-agnostic and could easily be repurposed for use with other NTDs.

## Data Availability

All data used in the manuscript are avaiable upon reasonable request to the authors.

